# Clinical and experimental factors that affect the reported performance characteristics of rapid testing for SARS-CoV-2

**DOI:** 10.1101/2021.05.20.21257181

**Authors:** Valentin Parvu, Devin S. Gary, Joseph Mann, Yu-Chih Lin, Dorsey Mills, Lauren Cooper, Jeffrey C. Andrews, Yukari C. Manabe, Andrew Pekosz, Charles K. Cooper

**Affiliations:** Becton, Dickinson and Company, BD Life Sciences – Integrated Diagnostic Solutions, 7 Loveton Circle, Sparks, MD, USA; W. Harry Feinstone Department of Molecular Microbiology and Immunology, Johns Hopkins Bloomberg School of Public Health; Department of Medicine, Johns Hopkins University School of Medicine, Baltimore, Maryland; Department of Emergency Medicine, Johns Hopkins University School of Medicine, Baltimore, Maryland

## Abstract

Tests that detect the presence of SARS-CoV-2 antigen in clinical specimens from the upper respiratory tract can provide a rapid means of COVID-19 disease diagnosis and help identify individuals that may be infectious and should isolate to prevent SARS-CoV-2 transmission. This systematic review assesses the diagnostic accuracy of SARS-CoV-2 antigen detection in COVID-19 symptomatic and asymptomatic individuals compared to RT-qPCR, and summarizes antigen test sensitivity using meta-regression. In total, 83 studies were included that compared SARS-CoV-2 rapid antigen lateral flow testing (RALFT) to RT-qPCR for SARS-CoV-2. Generally, the quality of the evaluated studies was inconsistent, nevertheless, the overall sensitivity for RALFT was determined to be 75.0% (95% confidence interval [CI]: 71.0-78.0). Additionally, RALFT sensitivity was found to be higher for symptomatic versus asymptomatic individuals and was higher for a symptomatic population within 7 days from symptom onset (DSO) compared to a population with extended days of symptoms. Viral load was found to be the most important factor for determining SARS-CoV-2 antigen test sensitivity. Other design factors, such as specimen storage and anatomical collection type, also affect the performance of RAFLT. RALFT and RT-qPCR testing both achieve high sensitivity when compared to SARS-CoV-2 viral culture.

## INTRODUCTION

Severe Acute Respiratory Coronavirus-2 (SARS-CoV-2) is the highly transmissible viral agent responsible for development of Coronavirus Disease 2019 (COVID-19).(Carlos et al., 2020;Chang et al., 2020;Li et al., 2020;Wang et al., 2020) Based on measurements from specimen swabs, the viral load in infected individuals peaks around the time of symptom onset (approximately 2-3 days following infection).(Walsh et al., 2020) This time point coincides with the highest rate of SARS-CoV-2 transmissibility. Transmissibility usually tapers off within 8 days following symptom onset.(He et al., 2020) Asymptomatic individuals account for 40-45% of all infections and can transmit the virus for up to 14 days following infection.(Oran and Topol, 2020) Therefore, rapid, accurate diagnostic testing has been a key component of the response to COVID-19, as identification of SARS-CoV-2-positive individuals facilitates both appropriate treatment and reduced communal spread of the virus.(La Marca et al., 2020)

Molecular testing using reverse transcription, quantitative polymerase chain reaction (RT-qPCR) platforms has become the primary diagnostic method for COVID-19 diagnosis.(Wang and Taubenberger, 2010;Tahamtan and Ardebili, 2020) The major advantage of RT-qPCR testing is its high analytical sensitivity (translating to few false negative results).(Giri et al., 2021) However, large-scale clinical laboratory testing requires a dedicated infrastructure, and specialized technician training. In addition, due to the specimen transport and processing time, results for standard RT-qPCR can take days to obtain, depending on the catchment area and the demand for testing.(Bustin and Nolan, 2020)

Antigen-based testing involves the application of specific, SARS-CoV-2 antibodies (Figure 1) in several formats, including lateral flow immunofluorescent sandwich assays, chromatogenic digital immunoassay, lateral flow immunoassay with visual read, and microfluidic immunofluorescence assays.(Rezaei et al., 2020) Antigen testing for SARS-CoV-2 can be utilized either in conjunction with RT-qPCR as a first-line screening test, or utilized in decentralized health care settings in which RT-qPCR testing may not be conducive for rapid result turn-around.(Rezaei et al., 2020) Rapid antigen-based lateral flow testing (RALFT) for SARS-CoV-2, as with influenza, has been implemented globally to achieve rapid, accurate results for COVID-19 diagnosis.(Peeling et al., 2021) Although the majority of antigen-based tests share a common mechanism for detection of SARS-CoV-2 protein, the reported sensitivities of both Food and Drug Administration (FDA) Emergency Use Authorization (EUA)-approved and non-EUA-approved antigen-based tests have varied greatly in the literature.(Brümmer et al., 2021) Multiple meta-analyses and systematic reviews have reported large inter-study heterogeneity related to SARS-CoV-2 antigen-based testing.(Dinnes et al., 2020;Brümmer et al., 2021) Although reliable antigen test performance coincides with high specimen viral load,(Brümmer et al., 2021) study heterogeneity could impact our conclusions about antigen test performance. Factors that could affect overall antigen test performance include: <u>analytical sensitivity</u> (i.e. antibody/antigen binding affinity) of the assay, which likely varies for tests across manufacturers,(Mina et al., 2020) <u>biases</u> occurring from the study design (e.g. blinding, test order, etc.), the <u>study population</u> (e.g. symptomatic versus asymptomatic, days from symptom onset [DSO], etc.), the <u>anatomical collection site</u> (e.g. nasopharyngeal versus anterior nares), and <u>specimen storage conditions</u>.(Lijmer et al., 1999;Griffith et al., 2020;Accorsi et al., 2021)

**Figure 1.**
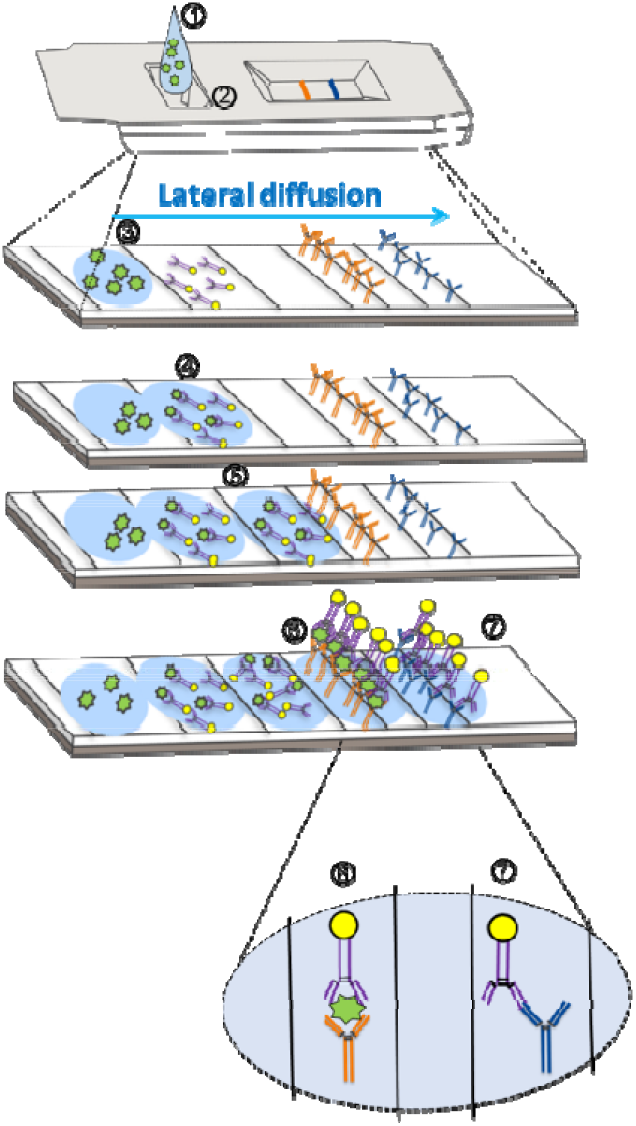
Mechanism of action for SARS-CoV-2 antigen detection through lateral flow assay design. **(1)** The specimen analyte, containing SARS-CoV-2 antigen suspended in assay buffer, is deposited in the sample well **(at 2)**. **(3)** The analyte (containing antigen; in green) is absorbed into the sample pad and begins to diffuse across the reaction chamber into the conjugate pad. **(4)** The analyte comes into proximity of a SARS-CoV-2-specific antigen antibody that is conjugated to a tag (usually consisting of gold, latex, or a fluorophore). **(5)**The antigen/antibody complex migrates via diffusion across the nitrocellulose membrane. **(6)** The SARS-CoV-2 antigen-antibody complex comes into proximity of a second SARS-CoV-2 antigen antibody (different epitope) that is covalently bound to the device pad and an antibody-antigen-antibody complex forms, resulting in the test line. Further diffusion of excess SARS-CoV-2 antibodies (unbound to antigen) results in association of a second, covalently-bound antibody that is specific for the first SARS-CoV-2 antibody. **(7)** An antibody-antibody complex forms resulting in the control line.

As others have noted previously, a wide range of reported sensitivities has been reported for rapid antigen testing.(Dinnes et al., 2020;Brümmer et al., 2021) The main objective of this meta-analysis was to explore possible causes of the high degree of heterogeneity of assay sensitivity estimates across different studies. Data were summarized and analyzed from over 80 articles and manufacturer IFUs to provide results on sensitivity for SARS-CoV-2 antigen testing from more than 25 individual assays.

## MATERIALS AND METHODS

The methods for conducting research and reporting results for systematic reviews and meta-analyses, which are outlined by the Cochrane Collaboration Diagnostic Test Accuracy Working Group and by PRISMA guidelines, were employed for this study.(Gatsonis and Paliwal, 2006;Leeflang, 2014;Page et al., 2020) This study protocol was registered with the PROSPERO International Prospective Register of Systematic Reviews in 2021 (PROSPERO CRD42021240421).(Booth et al., 2011)

The PICO (Participants, Intervention, Comparator, and Outcomes) of this meta-analysis was as follows: <u>P</u>articipants were individuals undergoing SARS-CoV-2 testing in a healthcare setting (at least 8 cases); Intervention (primary) was the index test consisting of a SARS-CoV-2 antigen detection platform utilizing immunobiological mechanisms, such as a sandwich ELISA, combined with spatial resolution (e.g. immunochromatographic assay); Intervention (secondary) was testing for SARS-CoV-2 using antigen and RT-qPCR testing (indices 1 and 2); <u>C</u>omparator (primary) was RT-qPCR as the reference test for detecting SARS-CoV-2 genomic RNA (any target gene); Comparator (secondary) was SARS-CoV-2 viral culture as the reference method for identifying specimens with infectious viral particles; <u>O</u>utcome was the determination of antigen test sensitivity across independent variables.

### Search and selection criteria

Eligible studies/sources included diagnostic studies of any design type (i.e. retrospective, prospective, randomized, blinded and non-blinded) that specifically involved the detection of SARS-CoV-2. The primary outcome was sensitivity for detection of SARS-CoV-2 in a healthcare setting by rapid antigen testing, as compared with RT-qPCR. Both MEDLINE and MedRxiv electronic databases were searched across dates ranging from January 01, 2020 to February 01, 2021 with the following search terms: (1) ((Antigen test and (sars-cov-2 OR COVID-19)) OR ((antigen[title/abstract] AND test) OR (Antigen[title/abstract] and assay)) AND (SARS-CoV-2[title/abstract] OR COVID-19[title/abstract])) and (2) “SARS-CoV-2 and antigen test or COVID-19 and antigen test”; respectively. In addition, a search was performed on the FDA database (https://www.fda.gov/medical-devices/coronavirus-disease-2019-covid-19-emergency-use-authorizations-medical-devices/vitro-diagnostics-euas#individual-antigen) for all EUA SARS-CoV-2 antigen tests. All retrieved sources were assessed for relevance using pre-determined inclusion/exclusion criteria. The inclusion criteria consisted of: (1) SARS-CoV-2 diagnostic target; (2) Sensitivity as a performance outcome; (3) Compares antigen testing performance with RT-qPCR as reference; (4) Population includes symptomatic and/or asymptomatic participants; (5) Human study; (6) English language; (7) Any region, country, or state. Secondary inclusion sub-criteria for analyses included: (S1) Index performance results that were stratified by viral load or by RT-qPCR (reference) cycle threshold; (S2) Delineated specimens for reference and index testing between symptomatic and asymptomatic participants; (S3) Delineated the anatomical site for specimen collection prior to reference and index testing; (S4) Delineated whether the specimen was frozen prior to reference and index testing; (S5) Specified whether the specimen was frozen prior to reference and index testing; (S6) Analytical limit of detection information was available for the reference assay; (S7) The index test manufacturer information was available. The exclusion criteria included: (1) Article/source from a non-credible source; (2) Article/source contains an unclear or indistinct research question; (3) Does not contain performance data specific to SARS-CoV-2; (4) Does not identify or does not involve standard upper respiratory SARS-CoV-2 specimens (e.g. contains other specimen types such as serological or saliva); (5) Contains no RT-qPCR reference results for comparison; (6) Data were collected in an unethical manner; (7) The index test involves a mechanism other than SARS-CoV-2 antigen detection involving a lateral flow (or similar) design; (8) Data not conducive for extraction required for analysis; (9) No data regarding true positive and false negative rates for the index test relative to the reference test. Additional, secondary exclusion criteria included (S1) Article/source not in the English language; and (S2) Study did not involve humans.

Full text reviews of the articles that passed initial screening were performed to identify those that met inclusion/exclusion criteria involving study methodologies, specimen collection, SARS-CoV-2 test details, and data type (sensitivity, specificity values, etc.) and format (raw data, only point estimates and 95% confidence intervals [95% CI] included, etc.), were identified and entered into data extraction tables to document study characteristics and to record raw data with calculated point estimates and 95% CI. A modified Newcastle-Ottawa Scale was used to evaluate the risk of bias (individual study quality),(Wells et al., 2011) which included the following bias domains: detection (measurement of test result), reporting (failure to adequately control confounding, failure to measure all known prognostic factors), and spectrum (eligibility criteria, forming the cohort, selection of participants). Risk of bias summary assessments for individual studies was categorized as “high”, “moderate”, or “low”. The overall quality of evidence for the risk estimate outcomes (all included studies) was obtained using a modified Grading of Recommendations, Assessment, Development and Evaluation (GRADE) (Schunemann et al., 2013) methodology for observational diagnostic studies.

The seven domains used to ascertain the overall study quality and strength across the six independent variables were (1) Confounder effect; (2) Consistency; (3) Directness; (4) Magnitude of effect; (5) Precision; (6) Publication bias; and (7) Risk of bias (ascertained from individual studies). Study sub-groups were considered high quality when ≥4 of seven domains received a green rating, with no red ratings and <3 unclear ratings; otherwise, it was considered moderate quality. Study sub-groups were considered moderate quality when three domains were green with <3 red domains; or when two domains were green and <3 domains were red with <4 domains unclear; or when 1 domain was green with <2 red domains and <3 domains were unclear; or when no domains were green, no domains were red and <2 domains were unclear. Any other combination of ratings resulted in a classification of quality as low

Subgroup meta-analysis was performed for the following factors: (1) viral load with fixed cutoff values; (2) symptomatic versus asymptomatic; (3) ≤7 DSO versus any DSO; (4) Anatomical collection type for specimens used for both index and reference testing (anterior nares/mid-turbinate versus nasopharyngeal/oropharyngeal); (5) Specimen storage conditions (fresh versus frozen); (6) Analytical sensitivity of the reference RT-qPCR test (detection cutoff <500 genomic copies/mL [cpm] versus ≥500 cpm); and (7) Assay manufacturer.

### Data analysis

Data extraction was accomplished by two reviewers/authors with any discrepancies adjudicated by a third reviewer/author. An independent author performed all statistical methods. All analyses were performed using R software (version 4.0.2) along with the meta (Balduzzi et al., 2019) and metaphor (Viechtbauer, 2010) packages. For each study, the sensitivity of the index test along with 95% Clopper-Pearson confidence intervals were calculated. Logit transformed sensitivity values were combined to obtain random effect estimates of overall sensitivity. The same method was applied to subgroup meta-analyses; subgroups were defined by disease status, reference and test collection type, reference and test storage (fresh / frozen), study spectrum bias, reference analytical sensitivity (high and low), and manufacturer. *Q-tests* for heterogeneity based on random effect models with common within-group variability were used to evaluate statistical differences between subgroups (univariate analysis). Moderators with significant heterogeneity in the subgroup analysis were included in a meta-regression mixed effect model. Forest plots were generated for all sub-group analyses; a funnel plot of all logit-transformed sensitivities was generated without taking into account study characteristics, and another funnel plot of residual values was generated after fitting the meta-regression model. Separately, for articles where viral load information was available, subgroup meta-analysis by viral load (either measured by RT-PCR Ct of 25 or 30, or a viral cpm of 1X10^5^) and symptomatic status was performed. The minimum number of studies required for synthesis is n=3.

## RESULTS

At the outset, 1,695 sources were identified during the database search (Figure 2). From that group of candidate sources, screening was performed by title and abstract, and the potential pool of articles was reduced, and 148 underwent full-text review for data extraction. Eighty-three (83) articles/sources of the 148 were chosen for meta-analysis (Table 1) based upon further exclusion criteria (see Materials and Methods). Note, a list of excluded sources with their specific exclusion criteria is available upon request from the authors. Of the 83 source studies, 76 (91.6%) involved a cross-sectional study design. Data from 12 of the sources (one of which is still pending authorization) were from validation studies as part of Emergency Use Authorization from the FDA and can be found in each, respective, manufacturer instructions for use. Twenty-two (22) of the studies were conducted in the USA, nine in Spain, seven each from Germany and Japan, six from Italy, four from China, three each from France, Switzerland, and the UK, two each from Belgium and Chile; the rest of the countries represented in this study had only one. One-hundred and thirty-five (135) individual data sets were utilized in total from the 83 articles/sources; 30 articles provided more than one data set. The overall, combined number of specimens from participants, from across all 83 studies, was 53,689; the overall total number of RT-qPCR reference positive results for estimating sensitivity from across all 135 data sets was 13,260.

**Figure 2.**
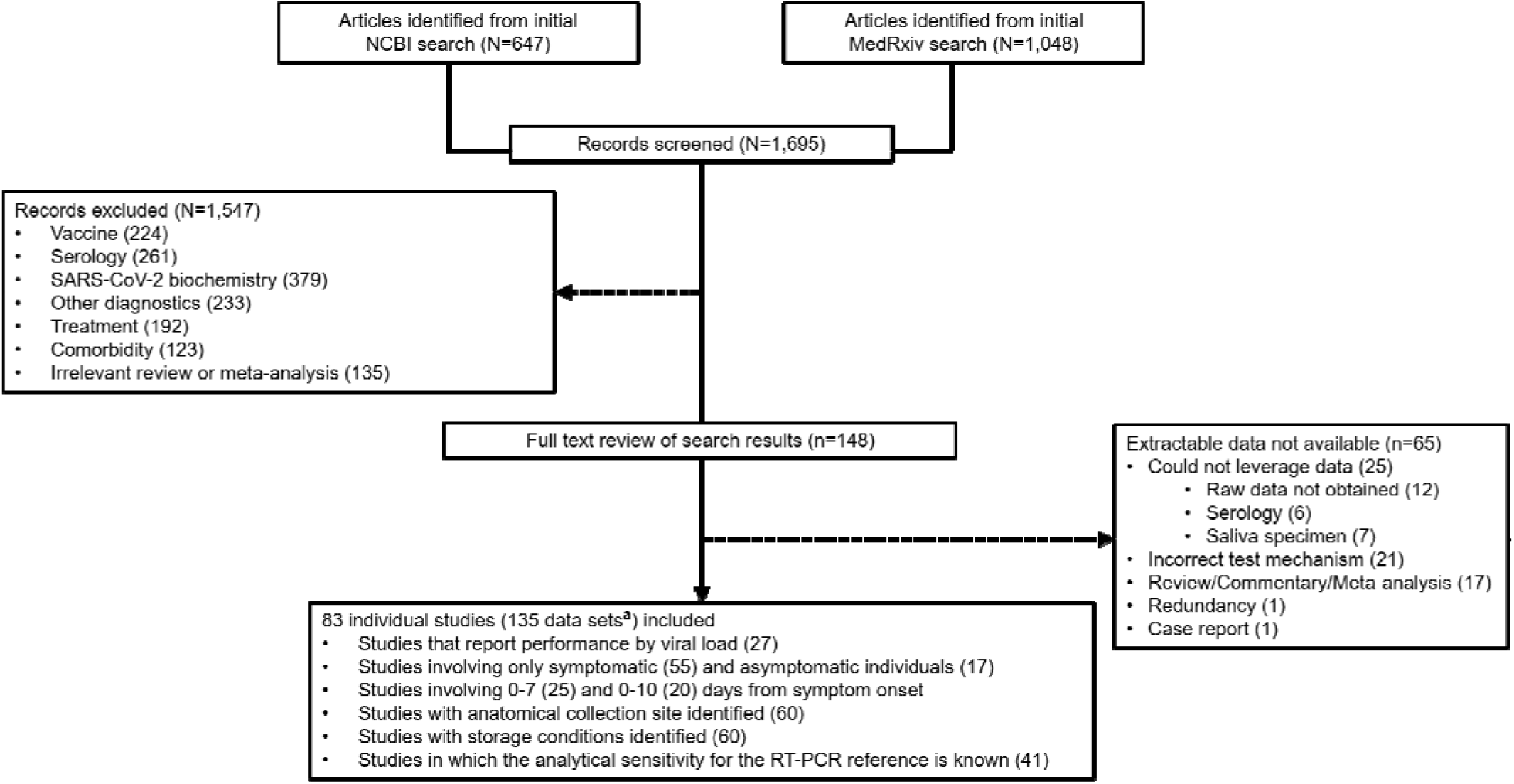
PRISMA flow diagram for reconciliation of articles/sources included in this study.

**Table 1.**
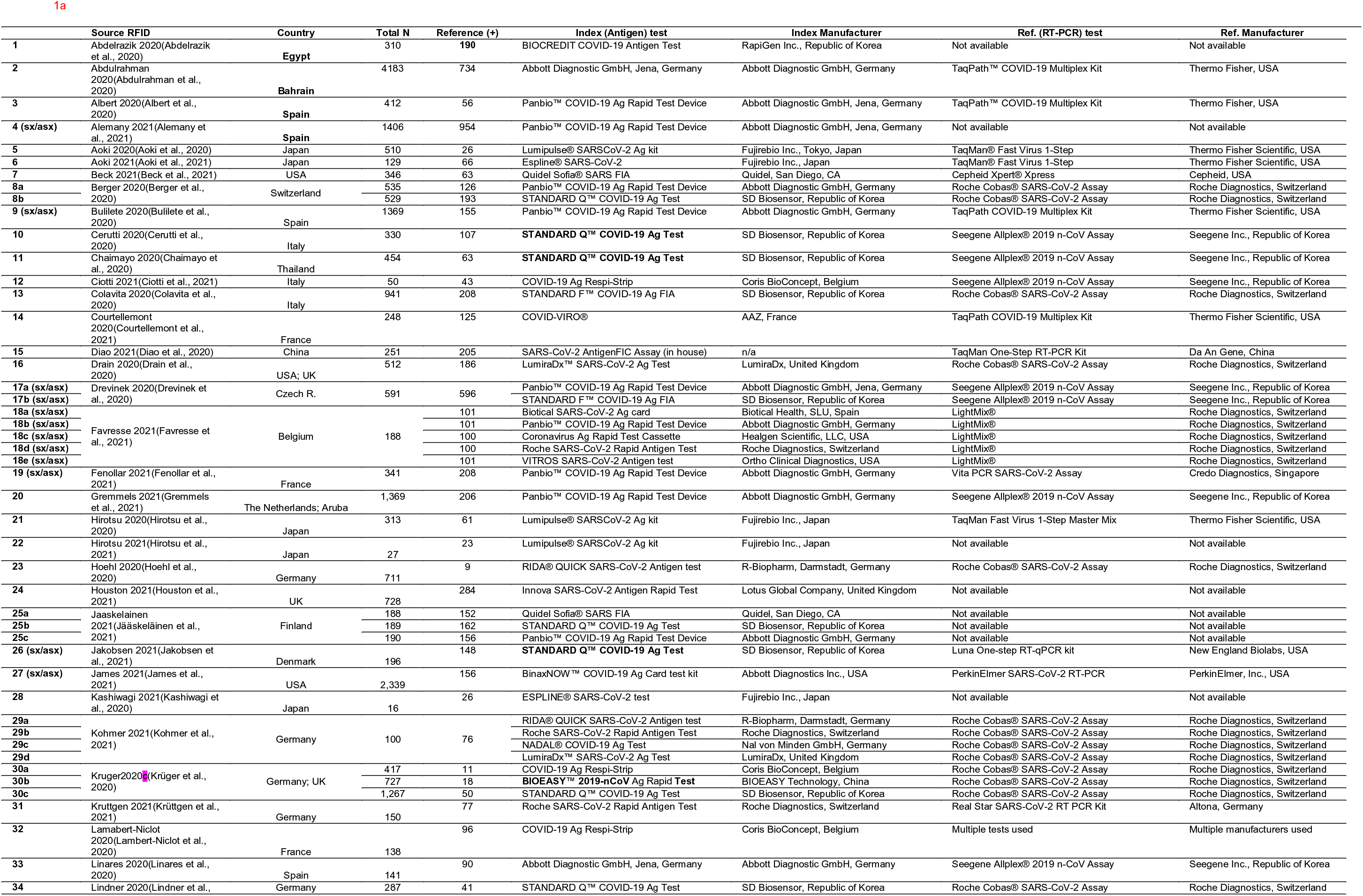
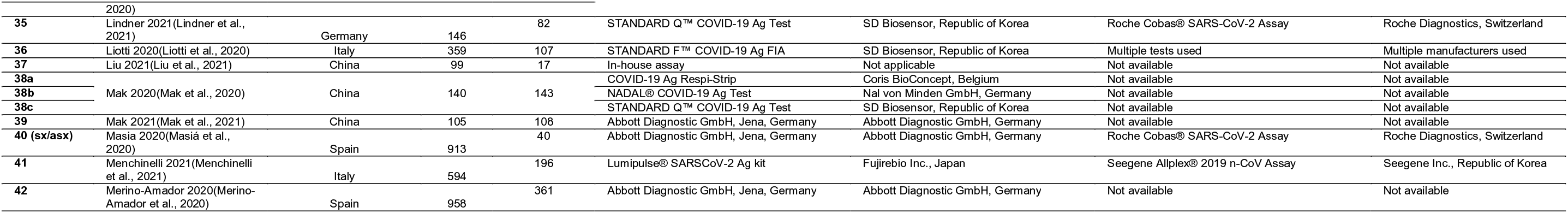
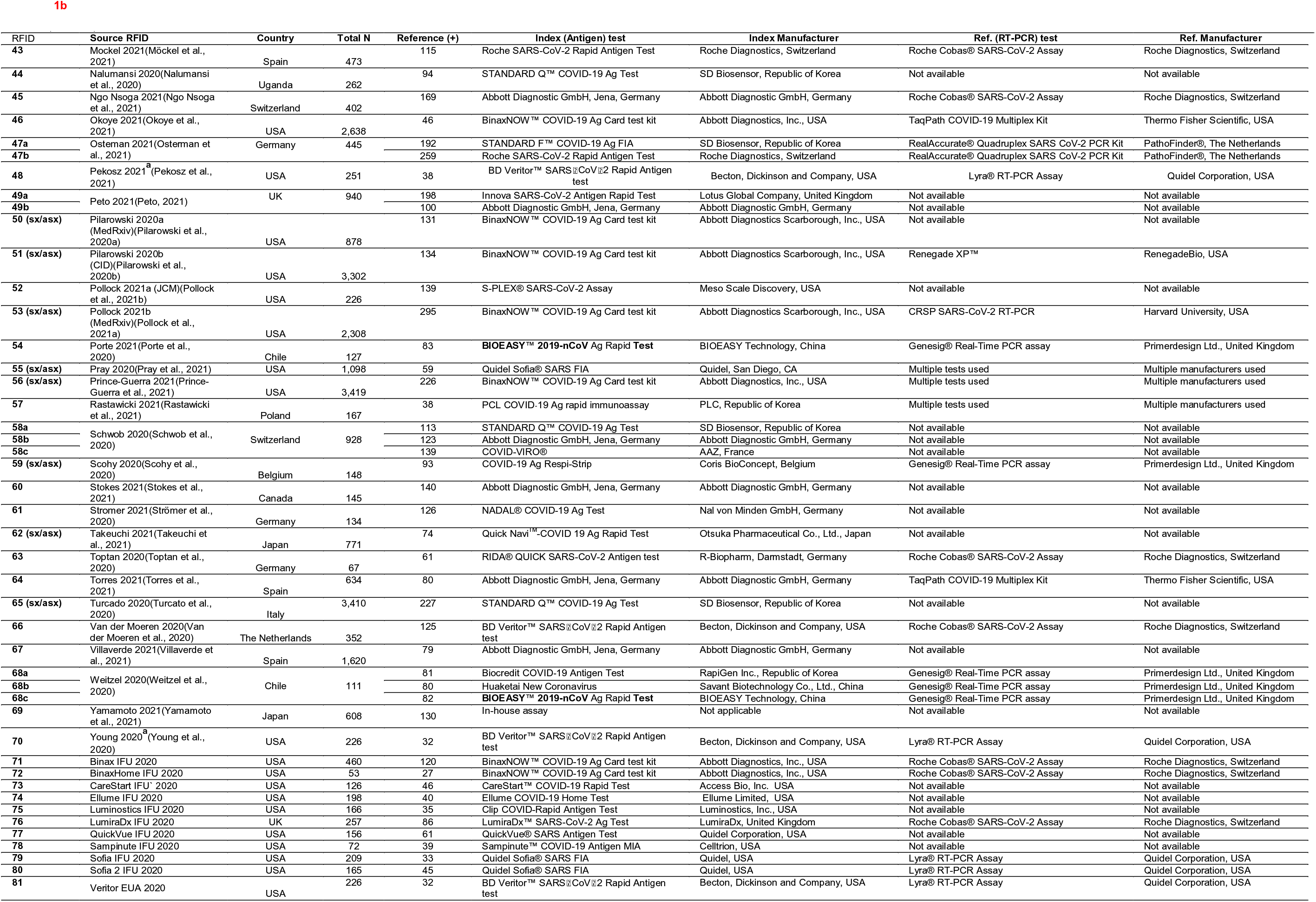
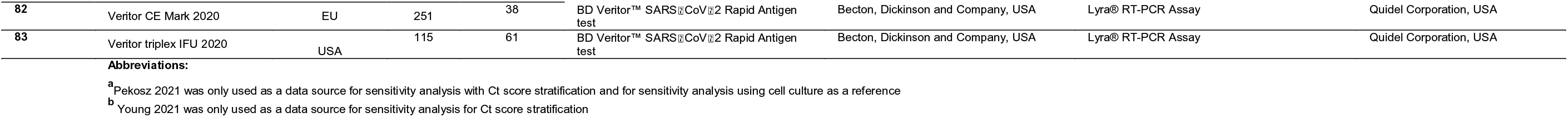
Information characterizing data sources involving SARS-CoV-2 antigen testing for analyses described in this report

A modified Newcastle-Ottawa Scale was used to rate biases of detection, performance, and participant selection (spectrum bias; Figure 3). The majority of studies were associated with low or moderate bias of detection (91.4%; 74/83), bias of performance (87.7%; 71/83), and spectrum bias (90.1%; 73/83). All included articles/sources had acceptable reference standards (which was an inclusion criteria), appropriate delay between index and reference testing, and no incorporation bias between the index and reference tests. The two most common weaknesses associated with study design for the included articles/sources were improper blinding and spectrum bias associated with participant enrollment.(Lijmer et al., 1999) Across the six primary factors (viral load, symptomatic versus asymptomatic, DSO, anatomical collection site, storage condition, analytical sensitivity of the reference RT-qPCR assay) analyzed here, the quality of evidence was quality of evidence was largely a mixture of high and moderate (Table 2).

**Figure 3.**
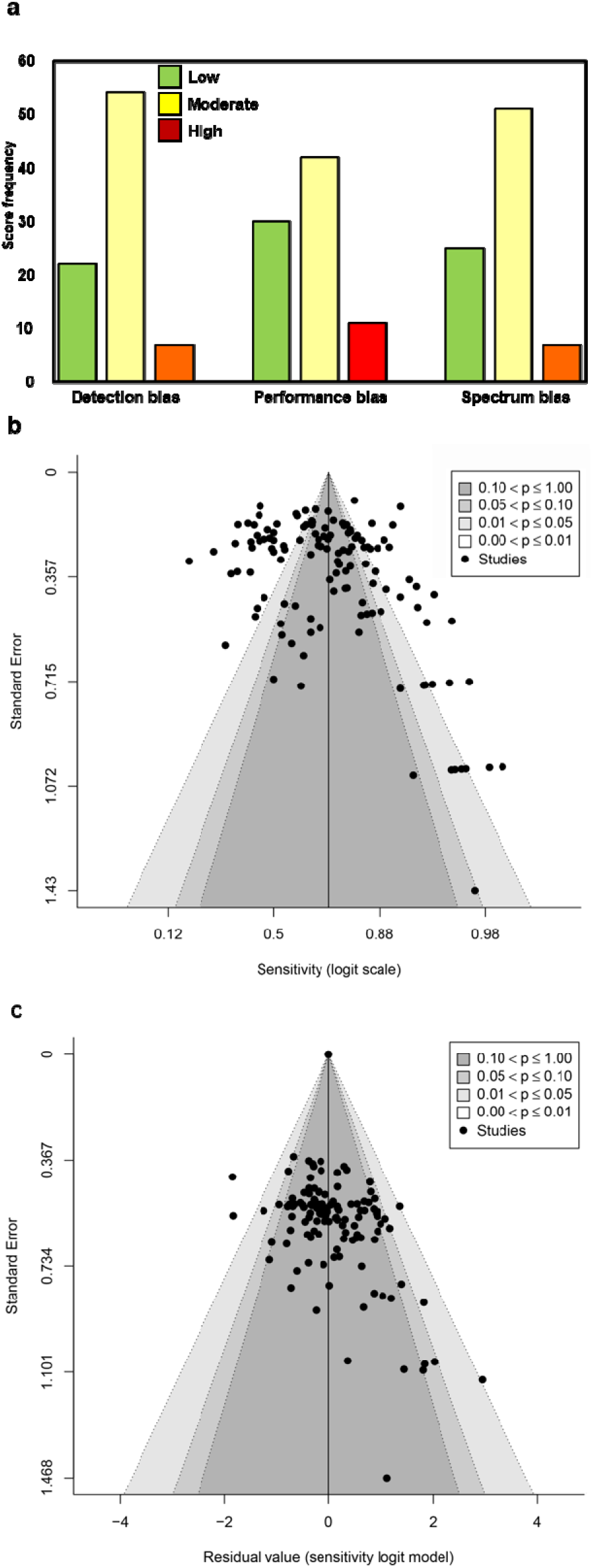
Determination of bias associated with the source articles/documents included in this meta-analysis. (a) Scoring as Low, Moderate, and High was performed for Detection bias, Performance bias, and Spectrum bias associated with each data source included. The frequency of the scores is plotted along the Y-axis. (b) and (c) Funnel plots of logit-transformed sensitivity in a model without any moderators (b) and with moderators included (c).

**Table 2.**
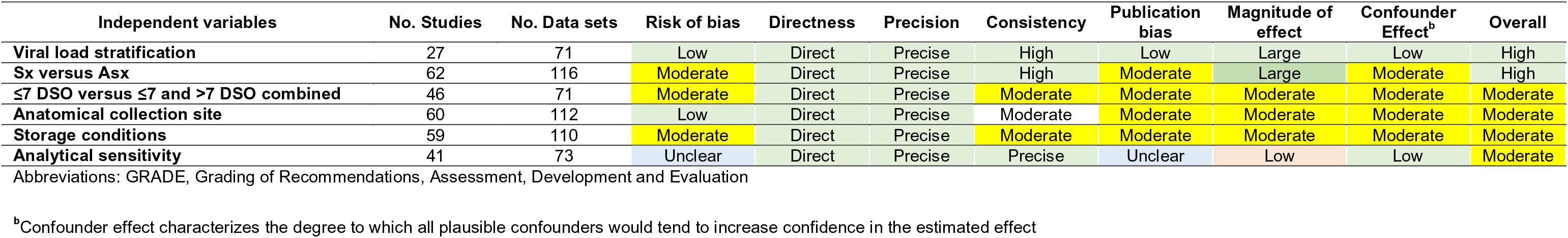
Overall quality of evidence for outcomes (modified GRADE) (Schunemann et al., 2013)

Eighty-three percent (83.0%; 112/135) and 16.3% (22/135) of the data sets provided data from COVID-19 symptomatic and asymptomatic individuals, respectively. The index test sensitivity point estimate (with 95% CI) for the symptomatic group (80.1% [95%CI: 76.0, 83.7]; reference positive n=9,351) was significantly greater (p-value < 0.001) than that for index test sensitivity for the asymptomatic group (54.8% [95%CI: 48.6, 60.8]; reference positive n=1,723). Of the 112 symptomatic data sets, 37.5% (42) included participants that were ≤7 DSO and 25.9% (29) included individuals that were a mix of ≤7 DSO and >7 DSO; 36.6% (41) had a DSO status that was unknown. A significant difference (p-value = 0.001) was observed when studies reporting on symptomatic individuals were sub-grouped by DSO; a sensitivity point estimate of 86.2% [95% CI: 81.8, 89.7] for the ≤7 DSO sub-group (reference positive n=3,480), compared to 70.8% [95% CI: 60.7, 79.2] for the group including both ≤7 DSO and >7 DSO (reference positive n=2,649) (Figure 4 and Table 3).

**Figure 4.**
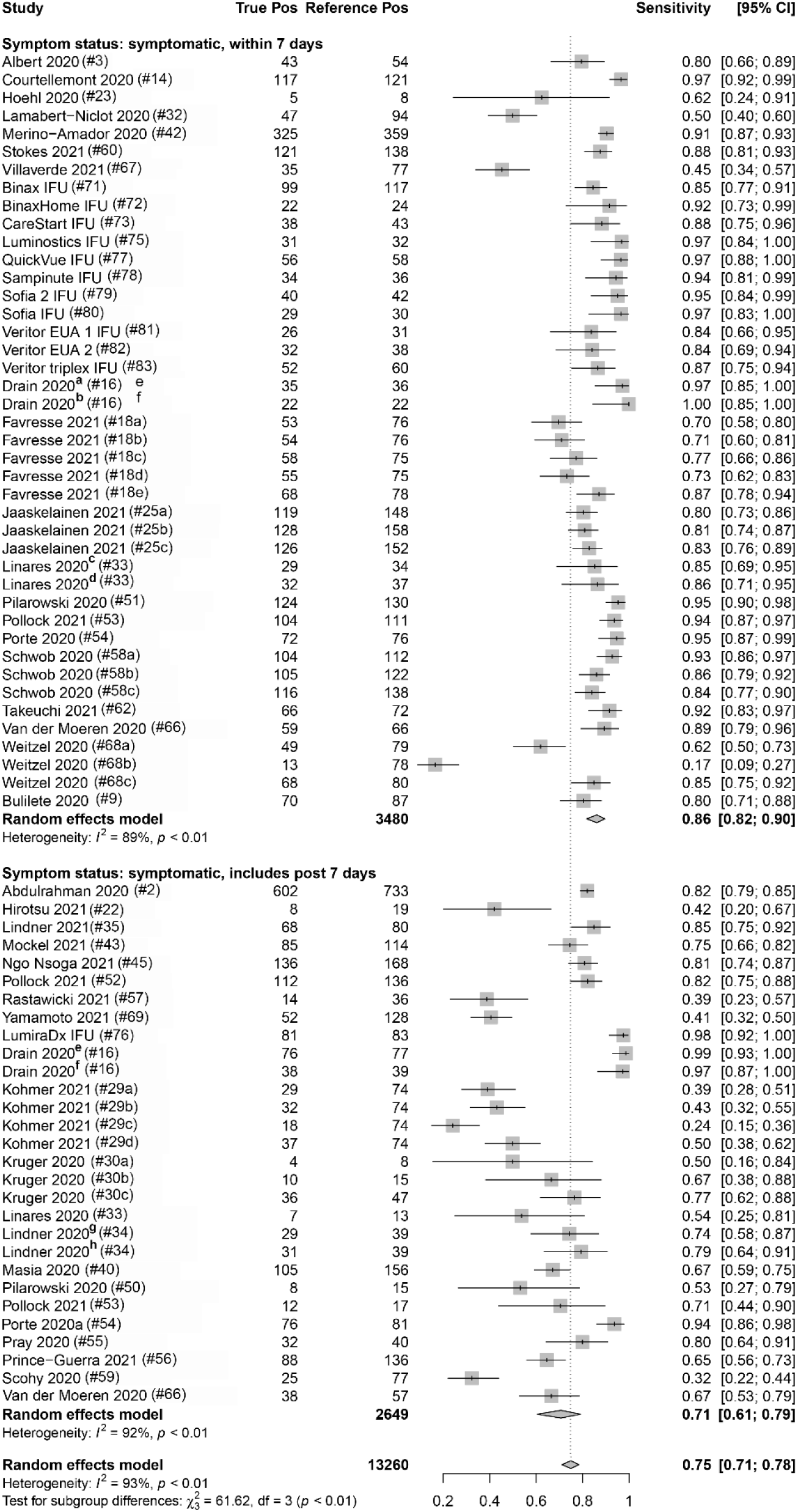
Forest plots containing calculated sensitivity values for index (SARS-CoV-2 antigen test) testing compared to reference (SARS-CoV-2 RT-qPCR assay) test. Data are stratified by days from symptom onset.

**Table 3.** Diagnostic performance (sensitivity) for antigen testing, with RT-qPCR as reference, stratified by different population and experimental factors

| Variable category | Variable group | No. data sets | Total Reference (+) | Sensitivity (%) | 95% CI | Univariate<sup>a</sup> | Multivariate |
| --- | --- | --- | --- | --- | --- | --- | --- |
| RT-qPCR Ct value | ≥10<sup>5</sup> cpm | 18 | 1,278 | **93.8** | [87.1, 97.1] | p<0.001 | n/a |
|  | <10<sup>5</sup> cpm | 18 | 781 | **28.6** | [16.2, 45.3] |  |  |
|  | Ct value ≤25 | 16 | 897 | **96.4** | [94.3, 97.7] | p<0.001 | n/a |
|  | Ct value >25 | 16 | 673 | **44.9** | [33.0, 57.4] |  |  |
|  | Ct value ≤30 | 37 | 2,536 | **89.5** | [85.3, 92.5] | p<0.001 | n/a |
|  | Ct value >30 | 37 | 679 | **18.7** | [12.9, 26.3] |  |  |
| Symptom status | Symptomatic | 95 | 9,351 | **80.1** | [76.0, 83.7] | p<0.001 | p < 0.001 |
|  | Asymptomatic | 23 | 1,723 | **54.8** | [48.6, 60.8] |  |  |
|  | Missing Information | 15 | 2,186 | **65.3** | [56.4, 73.3] |  |  |
|  | ≤7 DSO | 42 | 3,480 | **86.2** | [81.8, 89.7] |  |  |
|  | ≤7 DSO + >7DSO | 29 | 2,649 | **70.8** | [60.7, 79.2] |  |  |
| Anatomical site (ref.) | Nasal swab | 25 | 1,654 | **82.7** | [74.7, 88.5] | p= 0.037 | p = 0.023 |
|  | NPS | 97 | 10,607 | **73.1** | [68.5, 77.2] |  |  |
|  | Missing | 11 | 999 | **71.6** | [55.7, 83.6] |  |  |
| Specimens storage (index) | Fresh | 101 | 9,666 | **75.3** | [70.8, 79.3] | p = 0.375 |  |
|  | Frozen | 23 | 3,307 | **70.9** | [61.0, 79.1] |  |  |
|  | Missing | 9 | 287 | **81.5** | [69.3, 89.5] |  |  |
| Analytical sensitivity (Ref.) | High (≤500 cpm)) | 53 | 4,531 | **74.2** | [66.9, 80.4] | p=0.650 |  |
|  | Low (>500 cpm) | 22 | 2,582 | **76.8** | [67.2, 84.2] |  |  |
|  | Missing | 58 | 6,147 | **75.0** | [69.8, 79.5] |  |  |
| Study spectrum bias | High/Moderate | 52 | 5,524 | **81.4** | [75.5, 86.1] | p = 0.004 | p = 0.757 |
|  | Low | 81 | 5,871 | **70.4** | [65.4, 74.9] |  |  |
| All sources combined | n/a | 133 | 13,260 | **75.0** | [71.0, 78.0] |  |  |

**Abbreviations:** CI, confidence interval; DSO, days from symptom onset; Ct, cycle threshold; cpm, genomic copies per mL; NPS, nasopharyngeal swab;
<sup>a</sup>Q-test p-value for heterogeneity among subgroups calculated from random effects meta-analysis

Eighteen (18) data sets reported true positives and false negatives by viral load in the specimen; 37 and 16 data sets reported values by a Ct value of 30 and 25, respectively, for the RT-qPCR (reference) assay. When data were stratified by ≥1X10^5^ cpm (n=1,278 reference positive results) versus <1X10^5^ cpm (n=781 reference positive results), a significant difference was observed (p < 0.001) between the sensitivity point estimates (93.8% [95%CI: 87.1, 97.1] and 28.6% [95% CI: 16.2, 45.3], respectively). Similar findings were associated with studies that were stratified by a Ct value of ≤30 (n=2,536 reference positive results) and >30 (n=679 reference positive results) (89.5% [95%CI: 85.3, 92.5] and 18.7% [95% CI: 12.9, 26.3], respectively) and those that were stratified by a Ct value of ≤25 (n=897 reference positive results) and >25 (n=673 reference positive results) (96.4% [95%CI: 94.3, 97.7] and 44.9% [95% CI: 33.0, 57.4], respectively) Within a given viral load category, there were no statistical differences between studies performed with symptomatic subjects versus studies performed with asymptomatic subjects; this was true regardless of the exact definition of viral load category: Ct of 25, Ct of 30, or genome copies per ml of 10^5^ (Figure 5 and Table 3).

**Figure 5.**
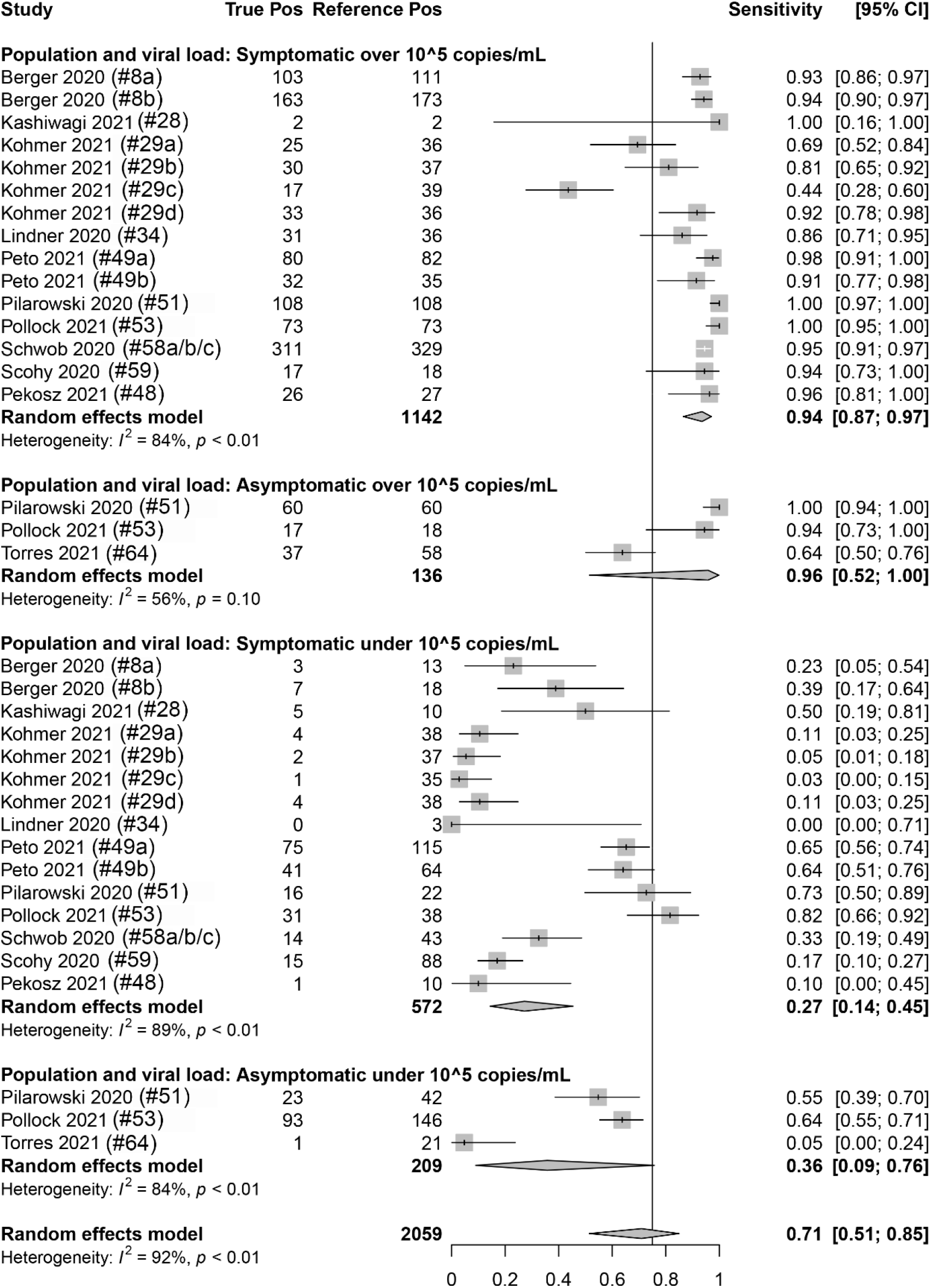
Forest plots containing calculated sensitivity values for index (SARS-CoV-2 antigen test) testing compared to reference (SARS-CoV-2 RT-qPCR assay) test. Data are stratified by viral load (genomic copies per mL; cpm) and by symptomatic or asymptomatic status.

True positives and false negatives by anatomic collection site were obtained from 97 data sets that included reference nasopharyngeal specimens and from 25 data sets that included nasal reference specimen. Antigen testing was usually paired from the same specimen type, only six datasets being non-paired (antigen nasal, reference nasopharyngeal) specimens. When analysis was performed on data stratified by anatomic collection site of the reference specimen, antigen test sensitivity was higher with a nasal specimen (82.7% [95%CI: 74.7, 88.5, p=0.037]; reference positive n=1,654) compared to a nasopharyngeal specimen (73.1% [95%CI: 68.5, 77.2]; reference positive n=10,607) (Table 3).

Storage condition of the collected specimens was also analyzed as one factor that could affect antigen test sensitivity. True positives and false negatives by test storage condition of the specimen that underwent antigen testing were obtained from 133 data sets. When analysis was focused on storage condition for index testing, antigen test sensitivity was 75.3% [95%CI: 70.8, 79.3] (reference positive n=9,666) for fresh specimens and 70.9% [95%CI: 61.0, 79.1] (reference positive n=3,307) for frozen specimens. This observed difference was not however statistically significant (p = 0.375) (Table 3).

Analytical sensitivity of the reference method (RT-qPCR) was determined using the manufacturer’s IFU when it was identified in the source documents and used to stratify true positive and false negative results associated with SARS-CoV-2 antigen testing. The LOD threshold for low and high analytical sensitivity was 500 cpm, which was the median (mean=582) LOD value for the analytical sensitivity from all of the reference methods included in this sub-analysis. Sensitivity values for antigen testing when stratified by high (reference positive n=4,468) and low (reference positive n=2,645) analytically sensitive reference method were similar: 74.2% [95%CI: 66.9, 80.4] and 76.8% [95%CI: 67.2, 84.2], respectively (Table 3).

Manufacturer (see Table S1) and study spectrum bias were also significant factors in subgroup meta-analyses; higher sensitivity was reported in studies with large/moderate spectrum bias (Table 3). A mixed-effects meta-regression model with moderators including symptom status, anatomical collection site, study selection/spectrum bias, and manufacturer was fit to the studies. All factors remained significant in the multivariate analysis, except study spectrum bias (multivariate p = 0.757). The moderators accounted for 72% of study heterogeneity (model *R^2^ = 0.722*). Visual inspection of unadjusted and multivariate-adjusted funnel plots for effect estimates from individual sources against study’s size was performed (Figure 3). The funnel plot asymmetry revealed possible reporting/publication bias reflecting fewer studies than expected that could be characterized by a small group number and a low sensitivity estimate for the index. Overall, study heterogeneity could largely be accounted for by the independent variables identified through sub-group analysis in this study.

### Culture as the reference

Sensitivity for SARS-CoV-2 antigen and RT-qPCR assays was determined as compared with SARS-CoV-2 viral culture as the reference method. There were five data sets that contained RT-qPCR (reference positive n=154) and antigen test (reference positive n=167) results. The overall sensitivity for RT-qPCR was 99.0% [95% CI: 96.0, 100] and for antigen testing was 90.0% [95% CI: 84.0, 94.0] (Figure 6).

**Figure 6.**
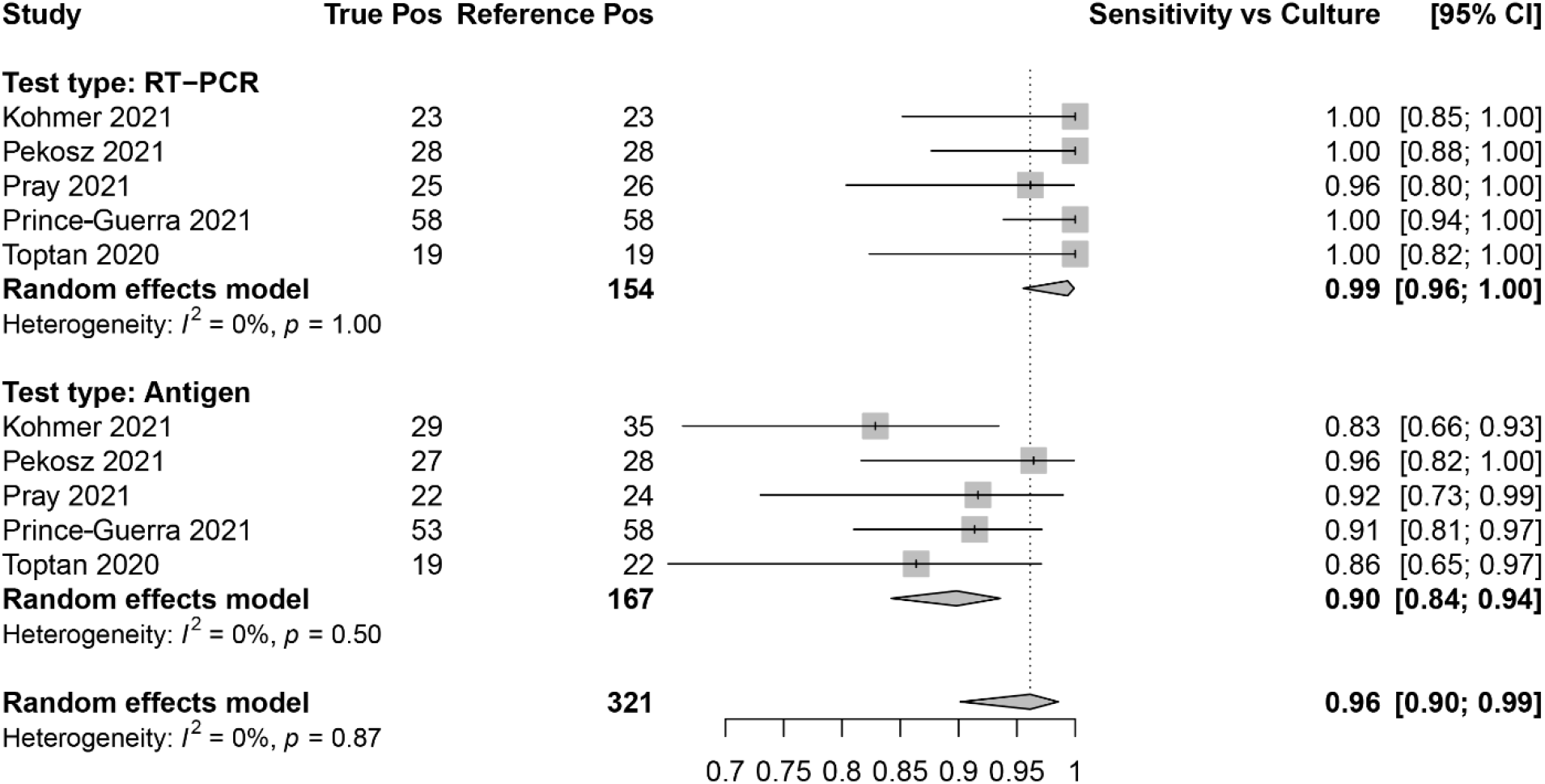
Forest plots containing calculated sensitivity values for index (SARS-CoV-2 antigen test and SARS-CoV-2 RT-qPCR assay) testing compared to reference (SARS-CoV-2 viral culture) test.

### Specificity

Raw data for false positive and true negative rates were extracted from 63 of 81 of the included studies; the overall specificity across the included studies was 99.4% [95%CI: 99.3, 99.4] for antigen testing compared to RT-qPCR as the reference.

## DISCUSSION

The PPA (sensitivity) point estimate for antigen testing, spanning the entire 135 data sets included here, was 75.0% (95% CI: 71.0, 79.0). We found that factors including specimen viral load, symptom presence, days from symptom onset, anatomical collection site, and the storage conditions for specimen collection could all affect the measured performance of SARS-CoV-2 antigen tests (Figure S1). In addition, our meta-analysis revealed that antigen test sensitivity (96.0% [95% CI: 90.0, 99.0]) was highest in SARS-CoV-2-positive individuals with an increased likelihood of being infectious at the time of testing (e.g., culture positive).{Pekosz, 2021 #8840} Although specificity data were not extracted for every study included in this meta-analysis, SARS-CoV-2 antigen testing had high specificity as published previously.(Brümmer et al., 2021) Experimental factors such as anatomical collection site, specimen storage conditions, analytical sensitivity of reference, and composition of the study population with respect to symptomology, all varied across the field of studies included here.

This meta-analysis adds to the conclusions of others that viral load is clearly the most important factor that influences sensitivity for SARS-CoV-2 antigen testing. Two related meta-analyses have been published to date. The first by Dinnes et al. (2020),(Dinnes et al., 2020) had the following key differences: (1) Dinnes et al. utilized numerous categories of POC tests beyond antigen based testing; (2) Dinnes et al. included five articles for antigen testing in their work; (3) Dinnes et al. did not stratify the meta-analysis results for antigen testing by study design/viral load as is performed in this work. The second by Brummer et al. (2021),(Brümmer et al., 2021) had the following key differences: (1) Brummer et al. focused on commercial rapid antigen tests; (2) Brummer et al. only included 48 articles for antigen testing in their work; (3) Brummer et al. did not stratify the meta-analysis results for antigen test performance by study design characteristics as is performed in this work.(Brümmer et al., 2021)

Test sensitivity was stratified by RT-qPCR Ct value (using both 25 cycles and 30 cycles as the cutoff), an inverse relationship was shown between Ct value and SARS-CoV-2 antigen test sensitivity. Both the ≤25 Ct group and the ≤30 Ct group had significantly higher sensitivities than their >25 Ct and >30 Ct counterparts, respectively, regardless of subjects’ symptom status. These results are consistent with those from previous studies. (Dinnes et al., 2020;Brümmer et al., 2021) However, Ct value has been shown by different groups to have low correlation between different RT-qPCR assays and platforms.(Ransom et al., 2020;Rhoads et al., 2020) RT-qPCR assays have different analytical sensitivities; a universal Ct value reference has not been established that can be used to define the optimal sensitivity/specificity characteristics for antigen testing. In addition to stratification by Ct value, analysis was also performed for SARS-CoV-2 antigen testing sensitivity by absolute viral load (using 1X10^5^ as the cutoff). When data were analyzed using this strategy, similar results were observed as for stratification by Ct value. The viral load threshold utilized here was determined by a consensus value that appeared with regular frequency from the source articles and represented a viral threshold that consistently delineated a zone, across which, the false positive rate increased for most antigen tests. It is generally accepted that viral loads of less than 1X10^5^ cpm correlate with non-culture positive levels. However, whether 1X10^5^ cpm is the most accurate threshold by which to measure antigen test performance is still a topic for debate. Some studies suggest that viral loads closer to 1X10^6^ cpm might be a more appropriate threshold, which would act to minimize false positive rates. (Berger et al., 2020;La Scola et al., 2020;Larremore et al., 2020;Quicke et al., 2020;van Kampen et al., 2020;Wolfel et al., 2020)

Several factors identified here that affect SARS-CoV-2 antigen test performance have been identified in previous studies to affect specimen viral load. For example, a significant difference in sensitivity for detection was noted here between symptomatic and asymptomatic individuals. However, stratification in both symptomatic and asymptomatic specimen groups by high viral load has a similar effect of increasing test sensitivity. Our data show that 82% of SARS-CoV-2-positive specimens from symptomatic individuals corresponded to a high viral load, whereas only 64% of SARS-CoV-2-positive specimens from asymptomatic individuals qualified as high viral load in this analysis. This bias may be due to the difficulty of estimating the timing of peak viral load in asymptomatic individuals when attempting to compare the natural history of viral load trajectory in symptomatic vs asymptomatic individuals.(Smith et al., 2021) Nevertheless, the presence of symptoms probably overlaps with higher specimen viral load, which subsequently affects the antigen test sensitivity. Anatomical collection type of the index and/or reference test method can affect the measured sensitivity estimates of antigen testing during a clinical trial; also, through a mechanism that involves increased/decreased viral load on the specimen swab. Evidence suggests that viral loads may be higher with nasopharyngeal than with nasal collection.(Pinninti et al., 2020) This difference may explain why measured antigen assay performance appears to be higher in studies that use a nasal RT-qPCR reference method.

Another factor identified here as potentially influencing measured antigen assay sensitivity was specimen storage, particularly with regard to the use of fresh vs frozen (i.e. “banked”) specimens. It is likely that protein antigen may, as the result of freeze/thawing, experience some degree of structural damage potentially leading to loss of epitope availability or a reduction in the affinity of epitope/paratope binding. Ninety-six (96) data sets involved fresh specimens for antigen testing and 23 data sets included freeze/thawed specimens for antigen testing. Although no statistically significant difference was detected between sensitivities for antigen test conducted on fresh versus frozen specimens, possibly due to the low data set group number in the frozen antigen group, a trend toward lower sensitivity was observed for tests performed on frozen specimens (75.3% [95% CI: 70.8, 79.3] for fresh versus 70.9% [95% CI: 61.0, 79.1] for frozen). In contrast, no similar trend was observed for specimen storage condition related to RT-qPCR testing (75.4% [95% CI: 70.6, 79.6] for fresh versus 77.7% [95% CI: 69.3, 84.3] for frozen). Additional results from in-house (i.e., a BD-IDS laboratory) testing with two different EUA authorized antigen assays demonstrate reduced immunoassay band intensity following freeze-thaw cycles, thus further supporting the findings from the meta-analysis that a freeze-thaw cycle could reduce analytical sensitivity for SARS-CoV-2 antigen testing (Figure S2).

The analytical sensitivity associated with the reference RT-qPCR assay was also investigated here as a possible variable that could affect the false negative rate of SARS-CoV-2 antigen testing. We hypothesized that relatively high analytical sensitivity for the reference RT-qPCR assay would impose a detection bias and result in decreased clinical sensitivity due to increased false negatives occurring near the RT-qPCR limit of detection. However, stratification by reference analytical sensitivity resulted in no difference in SARS-CoV-2 antigen test clinical sensitivity. It is likely that the analytical sensitivity of RT-qPCR, regardless of the manufacturer, is high enough that even relatively low sensitivity-RT-PCR assays are still well below the corresponding limit of detection for antigen testing. On the other hand, some manufacturers assay antigen test performance in a manner that involves sensitivity above and below a set Ct value. It is possible that analysis involving stratification by RT-qPCR analytical sensitivity could reveal differences in antigen test performance if all antigen test performances were determined in a similar manner that involves predetermined high/low viral load categories.

Several population and study design-specific factors were identified to be associated with higher measured assay sensitivity, likely due to the association with higher viral loads. This meta-analysis demonstrates that these factors exist in various combinations across studies in an inconsistent way, thus making comparisons of assay performance across these studies impossible. The lack of consistency across study designs makes it very difficult to compare point estimates between antigen tests to judge their relative clinical efficacy. The introduction of different forms of bias into study design, and during study conduct, could explain why discrepancies have been noted, for example, between sensitivity values listed in manufacturers’ IFUs and those obtained during independent evaluation of the same antigen test. Ultimately, direct comparison between antigen tests should be the most reliable approach for obtaining relative performance characteristics with any certainty. Here, we stratified SARS-CoV-2 antigen test sensitivity by spectrum bias associated with each of the data sources. We found that those studies rated with higher spectrum bias also had higher antigen test sensitivities. In addition, the funnel plot analysis that was performed for this meta-analysis shows obvious publication bias, which implicated a lack of publication of studies with low study group number and low sensitivity.

Clinical trials and studies involving diagnostics are vulnerable to the introduction of bias, which can alter test performance results and obstruct an accurate interpretation of clinical efficacy or safety. For example, antigen testing appears to have a higher sensitivity when compared to SARS-CoV-2 viral culture as the reference than when compared to RT-PCR. However, these two reference methods measure different targets: RNA only, versus infectious virus. Therefore, their use as a reference method should be intended to answer difference scientific questions, rather than artificially inflating apparent sensitivity point estimates. If the intent of a diagnostic test is determining increased risk of infectiousness through the presence of infectious virus, the high analytical sensitivity of RT-qPCR, which cannot distinguish RNA fragments from infectious virus, renders this diagnostic approach vulnerable to the generation of false positive results, particularly at later time points following symptom onset. At time points beyond one week from symptom onset, a positive RT-qPCR result more likely indicates that an individual has been infected, but is no longer contagious and cannot spread infectious virus. This is especially true for those with a SARS-CoV-2-negative cell culture result. Previous reports have shown that performance values for rapid antigen tests and SARS-CoV-2 viral culture exhibit better agreement than do results from RT-qPCR compared to viral culture.(Pekosz et al., 2021) This finding further supports the use of antigen testing for SARS-CoV-2 for the identification of individuals with infectious virus who are therefore at greater risk of virus transmission.

### Limitations

This study has some limitations. First, it was difficult to obtain reliable information across the sources, in a consistent manner, about disease severity, in order to perform meta-analysis on this aspect of COVID-19 diagnostics. Additionally, the studies included in this meta-analysis did not contain sufficient information to explore the potential effect of factors previously demonstrated to be associated with higher viral loads such as disease severity and community prevalence.

### Conclusion

In addition to viral load, several factors including symptom status, anatomical collection site, and spectrum bias all influenced the sensitivity for SARS-CoV-2 detection by antigen-based testing. This heterogeneity of factors found to influence measured assay sensitivity, across studies, precludes comparison of assay sensitivity from one study to another. Future consideration regarding standardization of these factors for antigen assay performance studies is warranted in order to aid in results interpretation and relative performance assessment.

## Data Availability

Data are available upon request.

## ACKNOWLEDGEMENTS

We thank Karen Eckert (Becton, Dickinson and Company, BD Life Sciences – Diagnostic Systems) for her input on the content of this manuscript and editorial assistance. The individuals acknowledged here have no additional funding or additional compensation to disclose.

## AUTHOR CONTRIBUTIONS

All authors contributed to the interpretation of the data, critically revised the manuscript for important intellectual content, approved the final version to be published, and agree to be accountable for all aspects of the work.

## SUPPLEMENTAL TABLES

**Table S1.** Antigen test sensitivity across brands (minimum number of studies = 3 to be included)

| **Antigen test name** | **No. data sets** | **Total Reference (+)** | **Sensitivity (%)** | **95% CI** |
| --- | --- | --- | --- | --- |
| Abbott Panbio | **28** | **4,400** | **74** | **[68, 80]** |
| BinaxNOW | 14 | 1,113 | **75** | [64, 84] |
| Lumipulse | **4** | **295** | **70** | **[48, 86]** |
| LumiraDx | 6 | 331 | **97** | [85, 99] |
| Respi-Strip | 6 | 372 | **37** | [30, 44] |
| RIDA Quick | 4 | 172 | **57** | [40, 73] |
| Roche RAT | 6 | 614 | **65** | [56, 73] |
| Sofia | 5 | 296 | **79** | [62, 90] |
| Standard F | 5 | 1,091 | **56** | [48, 65] |
| Standard Q | 15 | 1,427 | **80** | [72, 86] |
| Veritor | 5 | 252 | **83** | [75, 89] |

**Abbreviations:** CI, confidence interval; DSO, days from symptom onset; Ct, cycle threshold; cpm, genomic copies per mL; NPS, nasopharyngeal swab
<sup>a</sup>Q-test p-value for heterogeneity among subgroups calculated from random effects meta-analysis

## SUPPLEMENTAL FIGURES

**Figure S1.**
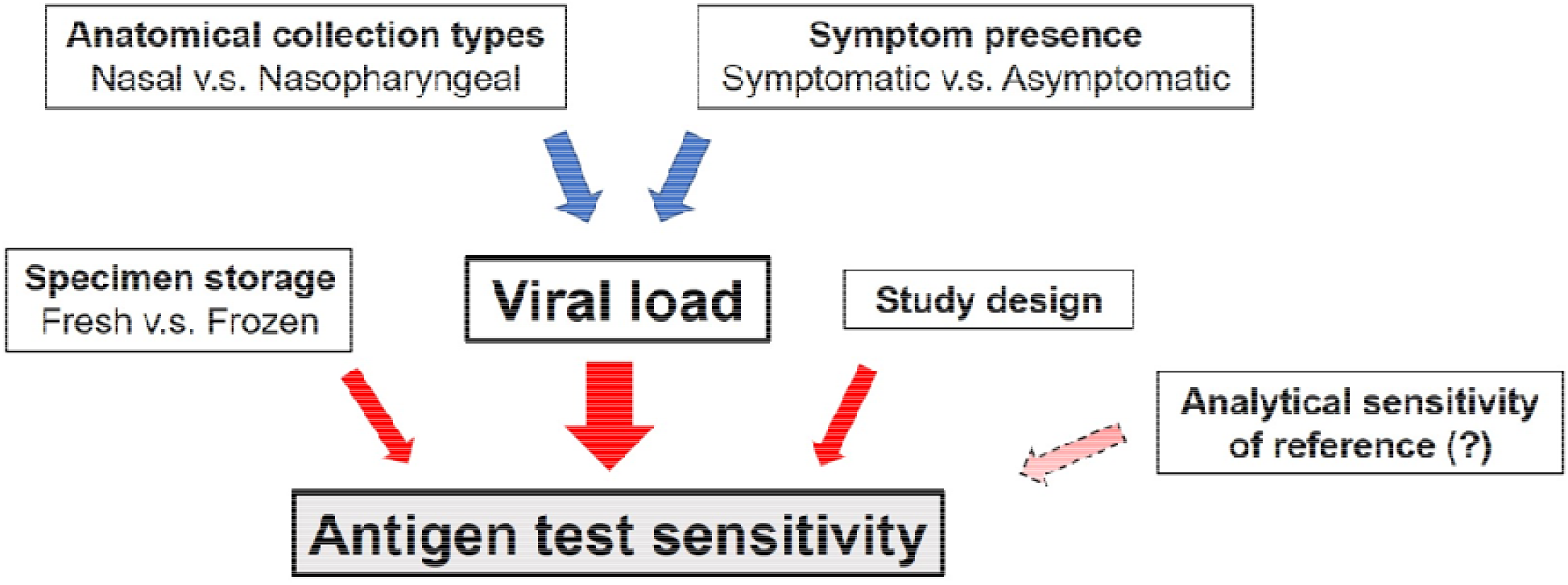
The relationship of factors that influence antigen test sensitivity

**Figure S2.**
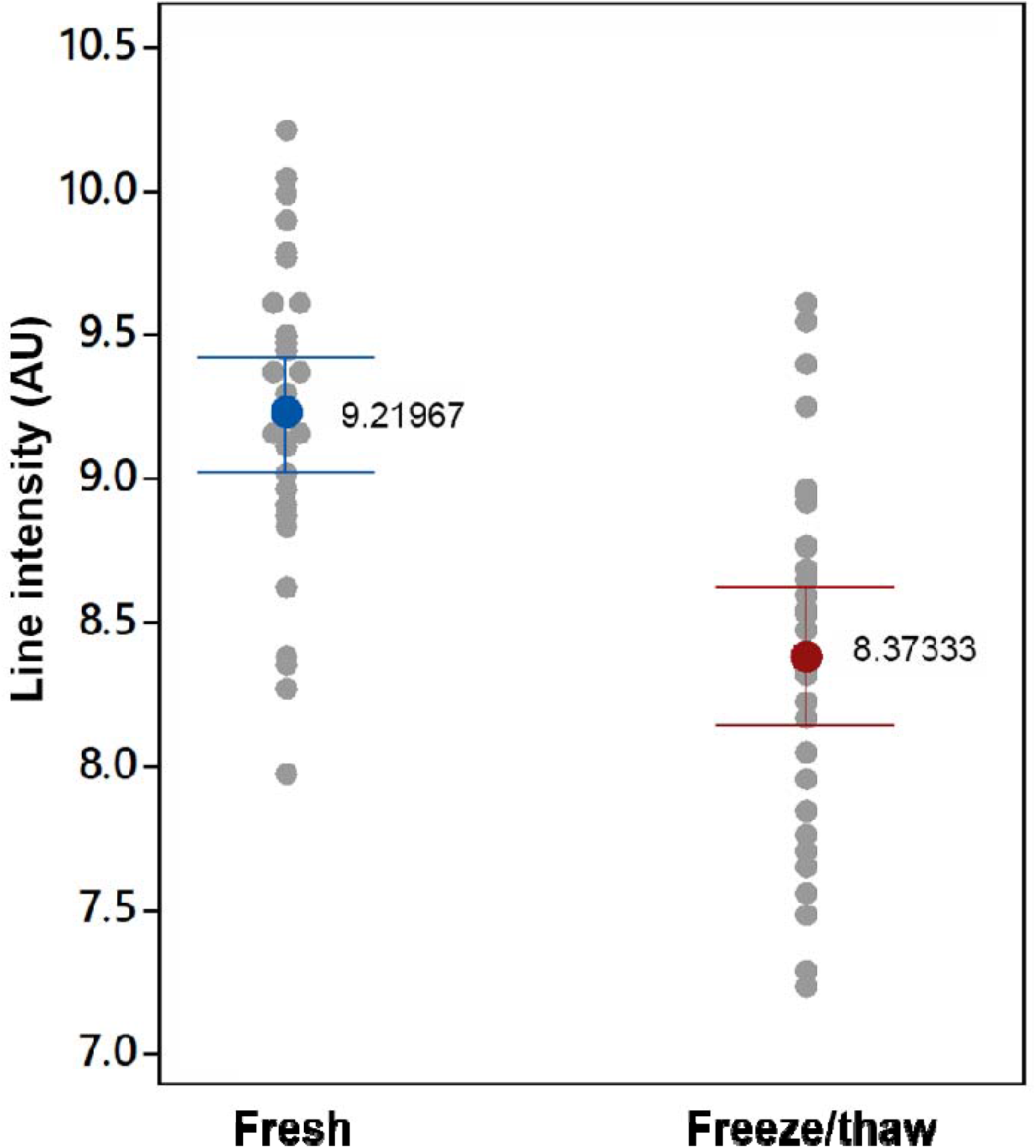
BD Veritor™ Plus Analyzer for SARS-CoV-2 antigen test Readings on RT-qPCR-positive SARS-CoV-2 specimens. Nasal specimens were collected and tested as described in the Manufacture’s IFU for the BD Veritor™ Plus Analyzer for SARS-CoV-2 antigen-test. One set of specimens was tested fresh and a parallel group was tested after one freeze/thaw cycle. All specimens were positive by the same RT-qPCR assay and there were no differences in the respective Ct values for the two antigen test groups.

